# *In-vitro* characterization of 2019-24 Influenza B Viruses reveals increased temperature-dependent fitness in later timepoints independent of antigenic drift

**DOI:** 10.1101/2025.10.27.25338757

**Authors:** Elgin Akin, Juanyu Zhang, David Villafuerte, Madeline Yunker, Anne P. Werner, Nicholas J Swanson, Richard E Rothman, Katherine JZ Fenstermacher, Heba Mostafa, Andrew Pekosz

**Affiliations:** Harry Feinstone Department of Molecular Microbiology and Immunology, Johns Hopkins Bloomberg School of Public Health, Baltimore, MD USA; Department of Pathology, Johns Hopkins University School of Medicine, Baltimore, Maryland, USA; Department of Emergency Medicine, Johns Hopkins University School of Medicine, Baltimore, Maryland, USA

## Abstract

Seasonal influenza viruses continually acquire mutations that can alter both their replication efficiency and antigenic properties. While recent seasonal Influenza B viruses (IBV) are characterized as a single clade, V1A.3a.2, defined by mutations in the hemagglutinin or HA protein, they have diversified globally into different subclades showing geographic restrictions. When the C.5 clade first emerged, it showed faster replication kinetics in human nasal epithelial cell (hNEC) cultures compared to clade A and C viruses from 2019-2021. The subsequent emergence of the C.5.1 subclade, predominantly observed in North and South America, resulted in no significant antigenic drift when compared to the 2023–2024 Northern Hemisphere vaccine strain, B/Austria/1359417/2021, using a vaccinated human serum set despite a number of mutations fixing in the HA protein. However, the C.5.1 subclade separated into several additional genotypes, some of which exhibit prolonged infectious virus production compared to the ancestral C.5 clade in hNEC cultures. C.5.1 viruses do not exhibit altered cell tropism hNEC cultures, indicating that differences in replication kinetics alone likely account for the observed increase in infectious virus production. The results demonstrate that IBV evolution can lead to changes in infectious virus production in the absence of antigenic drift.

**Importance:** Escape from preexisting population immunity or antigenic drift, is important to monitor with seasonal influenza viruses because it has an impact on vaccine strain selection. Less attention is paid to changes in virus replication which may impact disease severity or spread. Isolation and characterization of Influenza B viruses circulating since 2023 has shown that the emergence of several different V1A.3a.2 subclades has had little effect on sensitivity to vaccine induced immunity but has changed virus replication in human nasal epithelial cell cultures, suggesting that virus replication, rather than antigenic drift, was an important selective pressure on IBV evolution.

## Introduction

Seasonal influenza viruses evolve to evade pre-existing immunity and to gain a competitive advantage over other viruses in the circulating quasi-species (1). In addition to two subtypes of Influenza A viruses (H1N1 and H3N2), as of March 2020, one lineage of Influenza B virus (IBV) circulates causing seasonal flu epidemics. IBV causes a significant proportion of influenza-associated morbidity and mortality and is responsible for major disease burden in some years (2,3). After the disappearance of the B/Yamagata lineage, IBV activity of the B/Victoria lineage has returned to incidence levels similar to those seen before the COVID-19 pandemic (4–6).

The global population has been vaccinated against the B/Victoria V1A.3a.2 clade since its inclusion in the 2022–23 seasonal influenza vaccine (5). Influenza B clades are defined by genetic differences in the hemagglutinin (HA) gene, which shapes both antigenic identity and viral fitness (7). Mutations in HA and other genomic regions can modulate viral phenotypes, including receptor binding (8,9), replication efficiency (10–12), innate immune evasion (13–15), and transmissibility (16). Antigenic properties are determined by HA epitopes targeted by host antibodies, and substitutions within these sites can lead to antigenic drift, reducing antibody recognition (17). Continuous antigenic evolution necessitates frequent vaccine updates to maintain protection against circulating strains (18).

Since 2023, the V1A.3a.2 clade has diversified into multiple subclades, including C.5.1, C.5.6, and C.5.7. Mutations have also accumulated in other genes, including the neuraminidase (NA) which also evolves to evade population immunity through antigenic drift, but at a slower rate. Among these subclades, C.5.1, characterized by a lysine substitution at HA position 183 within the receptor-binding 190 helix, rose rapidly in frequency across North America and expanded into Europe and Asia. A sharp increase in C.5.1 detections at Johns Hopkins Health System toward the end of the 2023–24 season prompted further investigation into the phenotypic properties of this subclade (19).

Genotypic analysis revealed a clear temporal split within the C.5.1 population over the course of the season. Early C.5.1 viruses were defined by mutations at HA positions 229 and 189, and NA position 395 (I to V), but these variants were rapidly replaced by viruses bearing a distinct set of mutations, including HA 128K, 183K, and 202V. These substitutions localize to key structural domains of the HA head, including the 120 loop and the 190-helix, both of which contribute to receptor binding and are known antigenic sites (18,20,21). We sought to characterize the antigenic and replication properties of early and late C.5.1 viruses to better understand the effect of accumulated mutations on virus replication and antigenic structure.

## Results

### The emergence of the V1A.3a.2 subclade C.5.1 in the 2023-24 Season

Influenza B viruses co-circulated with Influenza A viruses in the geographical region served by the Johns Hopkins Health System during the 2023-24 season (**Figure 1A**), with IBV circulation increasing steadily both in total case numbers and positivity, showing near equivalent observed cases with IAV by April 2024.

**Figure 1.**
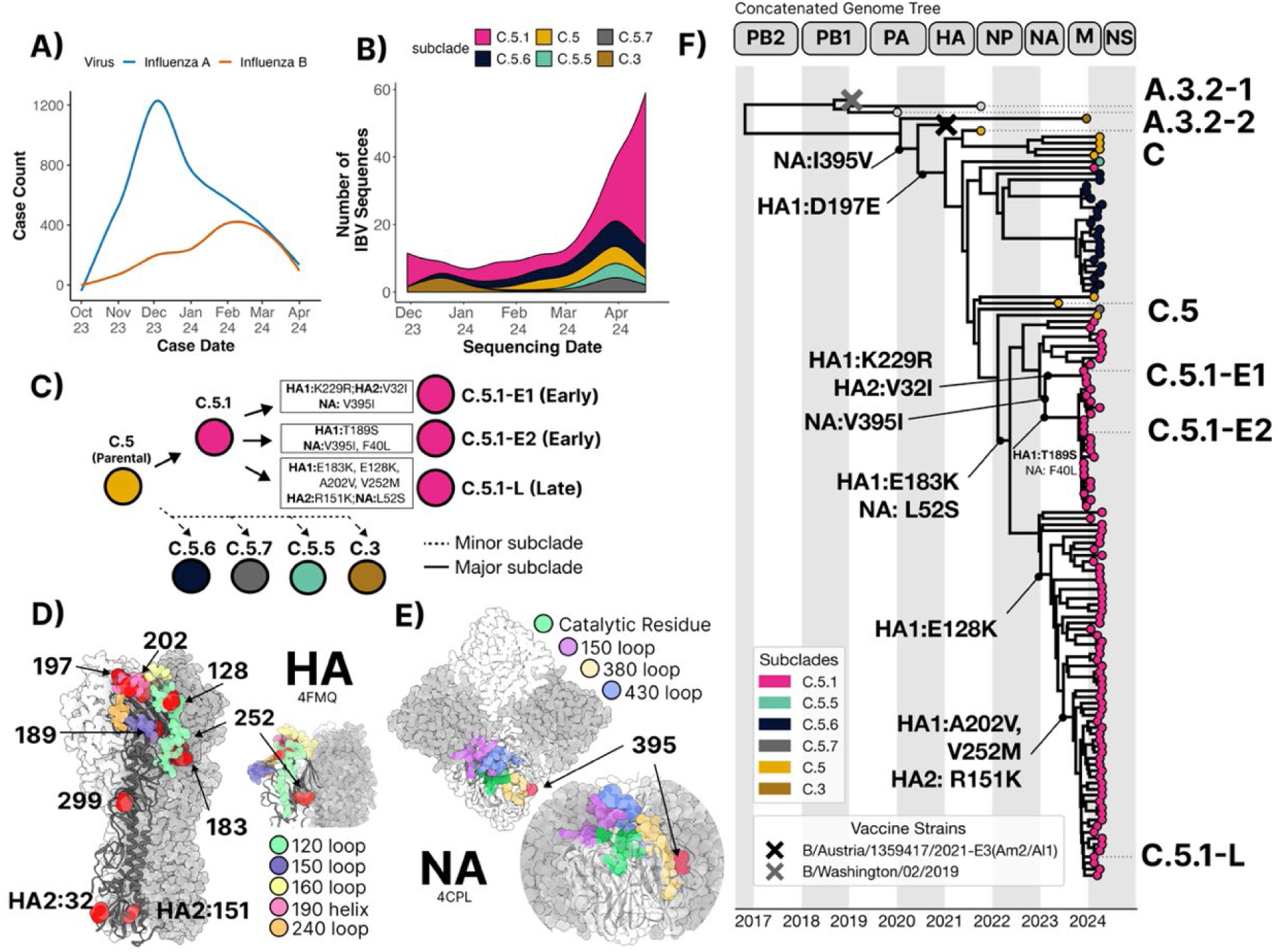
Phylodynamics of Influenza B viruses from Baltimore, MD in the 2023-24 Season. Hospital case counts **(A)** and HA subclade identities of sequenced specimens **(B)** of Influenza B/Victoria viruses presenting in the Johns Hopkins Hospital Network during the 2023-24 season. **(C)** A schematic of the V1A.3a.2 subclade diversification into the major C.5.1 with their denoted HA and NA mutations along with the minimally sampled C.5.6, C.5.7, C.5.5, and C.3 subclades. **(D)** Three-dimensional structures of the trimeric HA (PDB ID code: 4FMQ) and **(E)** the tetrameric NA (PDC ID code: 4CPL) proteins of B/Brisbane/60/2008 (Victoria lineage) mapped with C5.1-L mutations annotated in red. Known antigenic sites are colored in both structures. **(F)** Phylogenetic tree of segment concatenated genomes with sequencing date-resolved internal branches. Tree-tips are colored by HA subclade designation or vaccine identity. HA and NA ancestral branch mutations are annotated by coding sequence. An interactive build of the concatenated genome and each of the 8 individual segments is available at https://nextstrain.org/groups/PekoszLab-Public/akine/ibv2025/genome.

IBV genome sequencing was performed on 115 IBV positive nasal swabs and the data and subjected to phylogenetic analysis and reconstruction (22). All specimens belonged to the B/Victoria lineage V1A.3a.2 clade. Out of 115 HA sequences, 89 (77%) belonged to the C.5.1 subclade and 17 belonged to the C.5.6 subclade (15%) (**Figure 1B**). The remaining 8% of sequences belonged to the C.5, C.5.5, and C.5.7. Only 2 HA sequences (<3%) belonging to C.3 were observed. The C.5.1 HA and NA sequences obtained between October 2023, and March 2024 were genetically distinct when compared to C.5.1 specimen sequenced between March and April 2024 which we refer herein as C.5.1-Early (C.5.1-E) and C.5.1-Late C.5.1-L). C.5.1-E viruses carried HA1:T189S within the 150 helix (**Figure 1C** and **D**). C.5.1-L viruses acquired 5 additional HA mutations including E128K which falls within the 120 loop along with E183K and A202V within the 190-helix. C.5.1-L viruses also carry V252M which is both surface facing and adjacent to the 120-loop face. All 115 sequences carried 197E which falls within the 190-helix and RBS and has been implicated in antigenic drift events in previous severe IBV seasons (23,24) **and (Figure 1C** and **D**). The acquisition of the C.5.1-E mutations and purifying selection of HA1:189S and NA:395I (**Figure 1E**) was accompanied by a sudden surge in IBV case numbers commonly seen at the tail end of the influenza season. Because Early and Late C.5.1 viruses were genetically distinct at several critical antigenic sites on HA and NA, we selected 2 different C.5.1-E specimens (C.5.1-E1 and E2 in **Figure 1C-D**) along with 1 representative ‘late’ season specimen deemed C.5.1-L (**Figure 1C-D**). Phylogenetic reconstruction of concatenate IBV genomes revealed 2 major clusters of C.5.1 viruses with 2 minor clusters within the C.5.1-E outgroups, suggesting no reassortment events had occurred with these subclades in the 2023-24 season (**Figure 1F** and **Supplemental Figure 1**). Given the mutational load at HA and NA antigenic sites, we chose 2 representative C.5.1-E strains from each minor cluster and 1 C.5.1-L strain for further characterization of antigenicity and *in-vitro* fitness (**Table 1**). A single parental C. 5 strain was included as a common ancestor (**Table 1 and Table 2**).

**Table 1.** Representative viruses isolated between 2020-2024 for characterization against the 2022-23 **C.5 (**B/Baltimore/0546/2025 – bolded)

| short name | strain | type | NH season | clade | subclade | gisaid accession |
| --- | --- | --- | --- | --- | --- | --- |
| A.3.2-1 | B/Baltimore/R0696/2020 | circulating | 2019-2020 | V1A.3 | A.3.2 | EPI_ISL_17353886 |
| A.3.2-2 | B/Baltimore/JH-002/2021 | circulating | 2020-2021 | V1A.3 | A.3.2 | EPI_ISL_15763833 |
| C | B/Baltimore/JH-003/2021 | circulating | 2020-2021 | V1A.3a.2 | C | EPI_ISL_15763837 |
| C.5 | **B/Baltimore/0546/2023** | **circulating** | **2022-2023** | **V1A.3a.2** | **C.5** | **EPI_ISL_20209279** |
| C.5.1-E1 | B/Baltimore/JH-663/2023 | circulating | 2023-2024 | V1A.3a.2 | C.5.1 | EPI_ISL_18945546 |
| C.5.1-E2 | B/Baltimore/JH-665/2023 | circulating | 2023-2024 | V1A.3a.2 | C.5.1 | EPI_ISL_18945548 |
| C.5.1-L | B/Baltimore/JH-547/2024 | circulating | 2023-2024 | V1A.3a.2 | C.5.1 | EPI_ISL_19861034 |
| vaccine | B/Austria/1359417/2021 | 2023-24 Vaccine | 2022-23 | V1A.3a.2 | C | EPI_ISL_1519459 |

**Table 2.** Non-synonymous mutations between the 2023–2024 viruses chosen for characterization by V1A.3a.2 subclade designation. All mutations are relative to B/Brisbane/60/2008 positions. No non-synonymous mutations were detected in BM2 and is thus omitted from the table.

| V1A.2a.3 Subclade | Representative | Genome Mutation Load | HA1 | HA2 | NA | NB | PB2 | PB1 | PA | NP | M1 | BM2 | NS1 |
| --- | --- | --- | --- | --- | --- | --- | --- | --- | --- | --- | --- | --- | --- |
| C.5 | B/Baltimore/0546/2023 | 32 | A127T, R133G, P144L, N150K, K166X, G184E, N197E, K203R, R279K |  | P42Q, V71A, K343E, G378E, T395V, V401I | P42Q, V71A | A301T | M65I, N454D | E260G, K312R, S530T, F709V |  | T15I, R105K |  | R39K, E76K, Y104H, L130F |
| C.5.1-E1 | B/Baltimore/JH-663/2023 | 38 | A127T, R133G, P144L, N150K, K166X, E183K, G184E, T189S, N197E, K203R, R279K |  | F40L, P42Q, L52S, V71A, K343E, G378E, T395I, V401I | F40L, P42Q, L52S, V71A |  | M65I, N454D, I462V | K312R, S530T, F709V | M9I, T55A, S58N | T15I, R105K |  | Y104H, L130F |
| C.5.1-E2 | B/Baltimore/JH-665/2023 | 36 | A127T, R133G, P144L, N150K, K166X, E183K, G184E, N197E, K203R, R279K, K299R | V32I | P42Q, L52S, V71A, K343E, G378E, T395I, V401I | P42Q, L52S, V71A |  | M65I, N454D, I462V | K198R, K312R, S530T, F709V | M9I | T15I, R105K |  | Y104H, L130F |
| C.5.1-L | B/Baltimore/JH-547/2024 | 38 | A127T, E128K, R133G, P144L, N150K, K166X, E183K, G184E, N197E, A202V, K203R, V252M, R279K | R151K | P42Q, L52S, V71A, K343E, G378E, T395V, V401I | P42Q, L52S, V71A |  | M65I, N454D, I462V | K312R, S530T, F709V | M9I | T15I, R105K |  | Y104H, L130F, A212V, V263M |

### The parental C.5 subclade has reduced viral fitness compared to ancestral viruses collected between 2020-2022 in a time-point and temperature-dependent manner

Prior to the emergence of C.5.1, the entirety of North America experienced low Influenza A and B detection following the wake of interventions implemented early into the SARS-CoV-2 pandemic (4,22,25,26). We sought to understand the *in-vitro* fitness kinetics of viruses leading up to and following this bottleneck of Influenza B circulation. We selected representative viruses circulating strains between 2020-2022 from the V1A.3 clade (B/Washington/2/2019-like) and early emergent viruses from the V1A.3a.2 parental subclades C and C.3 which predate C.5.1.

The physiological relevance of polarized human nasal epithelial cell (hNEC) cultures can reveal virus fitness differences not apparent in widely used immortalized cell line models (8,10,11,27). Mutations previously observed in the 190 helix of HA1 have been shown to impact viral fitness which were present in the subclade C.5 strain but not in the parental subclade C and A.3.2 viruses has been shown to impact viral fitness (**Table 1 and Table 2**) (10). To determine whether the C.5 clade acquired a replication fitness advantage over ancestral strains, we compared the replication kinetics of a C.5 virus isolated in the 2022-23 season with isolates from the 2019-2020 and 2020-2021 Northern Hemisphere influenza seasons belonging to the parental V1A.3 subclade A.3.s and V1A.3a.2 subclade C (**Table 1 and Table 2**). Low MOI growth curves were performed on human nasal epithelial cell (hNEC) cultures at either 33°C, the upper respiratory tract temperature or 37°C the lower respiratory tract temperature. The 2022-23 C.5 representative produced significantly more infectious virus at 12-, 24-, and 36-hour post infection (hpi) before peak production at 48hpi, but between 48-120hpi, C.5 produced significantly less virus at both temperatures (**Figure 2A** and **E**). Area under the curve (AUC) analysis was used to calculate total virus production prior to and post 48 hours post infection (respective to the C.5 peak). C.5 produced significantly higher virus between 2-48hpi never reached the same peak viral titer compared to its parental strains at 33°C or 37°C (**Figure 2A-F**). When comparing replication at 33°C and 37°C, no temperature-specific differences were observed during early time points (12–48 hpi), except for A.3.3-2 (**Figure 2G–J**). However, after 72 hours, virus production was significantly lower at 37°C across all strains, regardless of subtype. These findings suggest that the C.5 subclade had a fitness advantage over prior IBV clades at early timepoints, but that was not observed at later time points.

**Figure 2.**
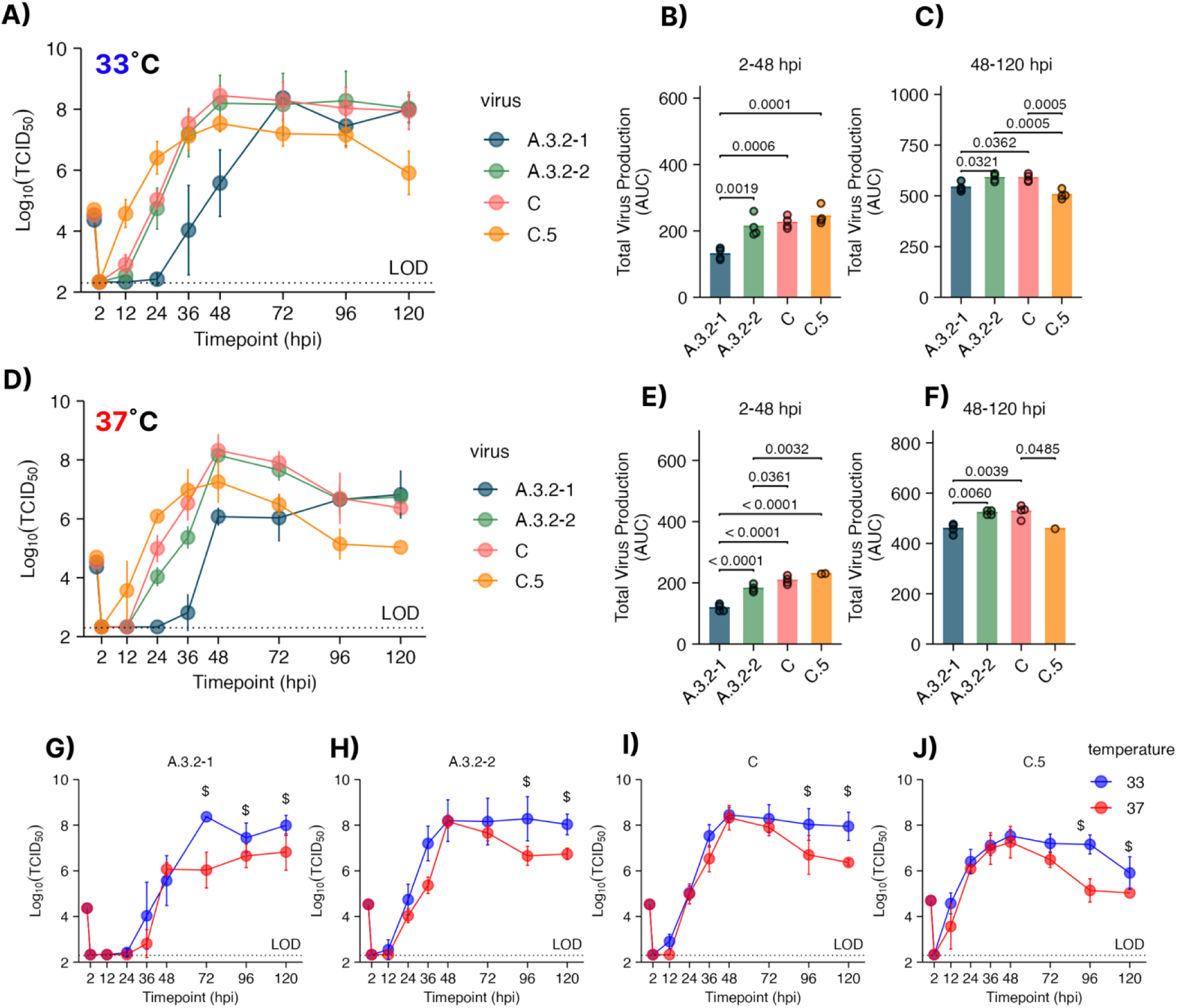
Characterization of Influenza B virus growth kinetics prior to the V1A.3a.2 C.5 subclade emergence. Low MOI Growth curves on human nasal epithelial cell (hNEC) cultures (MOI = 0.01) at 33°C **(A)** or 37°C **(D)** reveal significantly different kinetics in early and late time points. (B-C) Virus production at 33°C between **(B)** 2-48 hours post infection(hpi) or **(C)** 48-120 hpi. Virus production between at 37°C at **(E)** 2-48 hours post infection (hpi) or **(F)** 48-120 hpi. **(G-J)** Growth curves plotted by virus at 33°C and 37°C per panel. Statistical testing was performed by one way (AUC) or two-way (growth curves) ANOVA with Tukey post host test. Each graph combines two or three experiments with four hNEC wells per experiment.

### The C.5.1 subclade is antigenically matched to the parental C.5 subclade and 2023-24 NH Vaccine

Vaccinations in the 2023-25 Northern Hemisphere (NH) season were shown to offer protection against infection and hospitalization across representative Influenza B Strains collected throughout the season (28,29). Furthermore, ferret sera raised against B/Austria/1359417/2021 recognized representative V1A.3a.2 viruses by HI GMTs suggesting a well-matched selection to circulating Influenza B strains (19). Experiments using human sera are another critical component of antigenic characterization. To evaluate how V1A.3a.2 influenza B virus (IBV) subclades are recognized by human serum antibodies, neutralization assays were conducted using serum samples collected on day 0 and 28 days after vaccination from 77 individuals in the 2023–2024 Northern Hemisphere influenza vaccine cohort (**Figure 3A**). The C.5.1-E1, C.5.1-E2 and C.5.1-L subclades (**Table 1**) showed similar mean serum neutralizing antibody titers as those that recognized the vaccine subclade C strain (B/Austria/1359417/2021) and parental C.5 strain (B/Baltimore/0546/2023) at baseline (**Figure 3B**) and post vaccination (Figure 2C). No significant differences in the NT_50_ fold-change between timepoints were seen across both circulating subclades with comparable numbers of participants reaching seroconversion, defined as a NT_50_ fold change ≥4, between all 5 viruses (**Figure 3D**). For the vaccine virus, C.5, C.5.1-E2 and C.5.1-L, higher NT_50_ values were observed in females compared to males at D28 (**Supplemental Figure 2**). Furthermore, females were observed to have higher seroconversion rates than males across all 4 strains. There were similar numbers of sero-negative individuals across the vaccine, C.5 parental and circulating C. 5. 1. viruses (**Figure 3B-C**). Following vaccination, the number of seronegative individuals was similarly reduced across all tested viruses. Finally, the fold increase in serum neutralizing antibody titer between D0 and D28 ranged from 9.93 to 14.33 but did not differ between viruses (**Figure 3E**). Together, these data demonstrate that the parental C.5 and circulating C.5.1 viruses are well matched to the 2023-24 B/Austria/1359417/2021 vaccine component.

**Figure 3.**
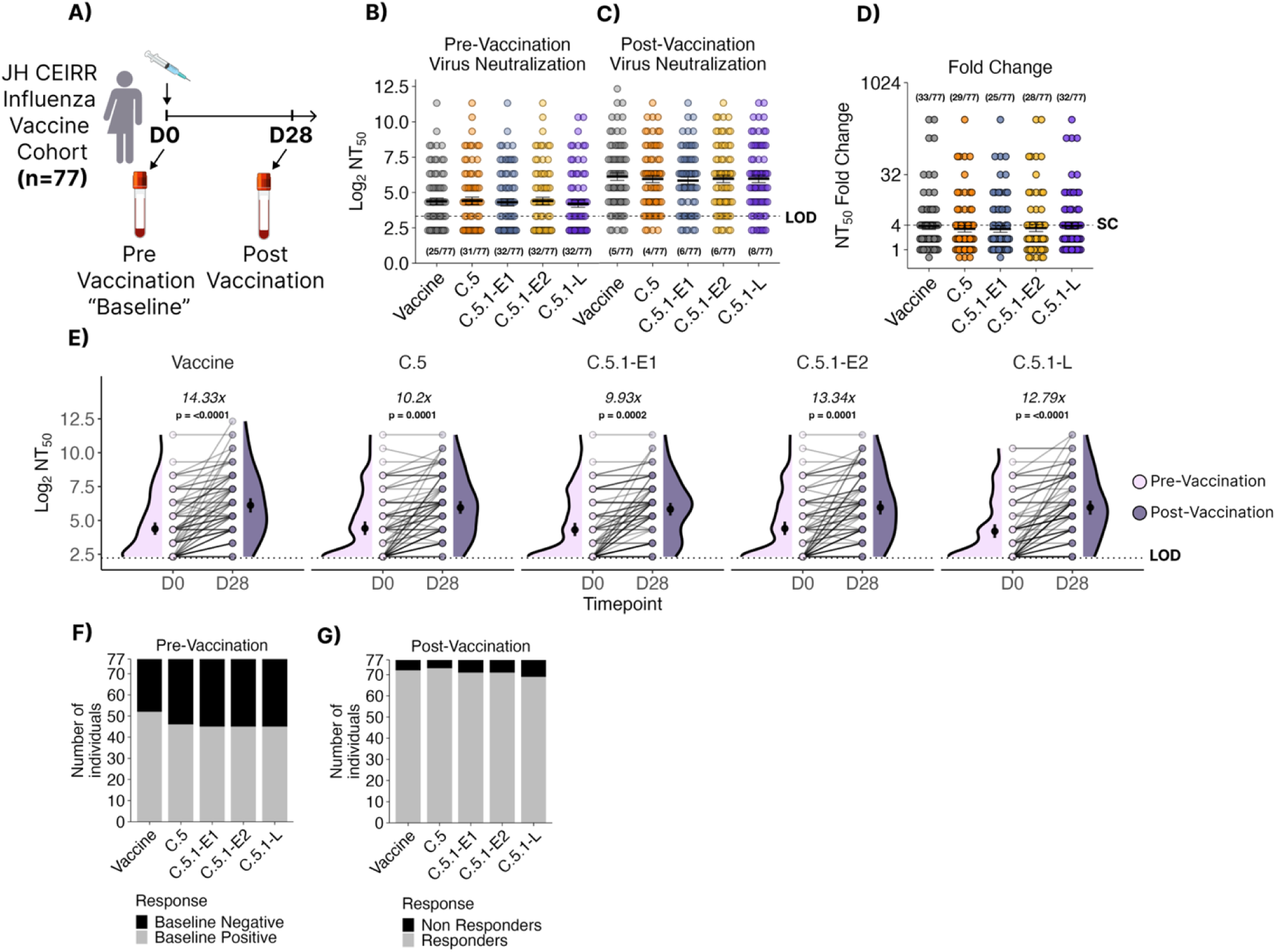
Neutralizing antibody titers (NT_50_) to in serum of healthcare workers is higher after vaccination. (A) The Johns Hopkins CEIRR Network influenza vaccine cohort consisted of 77 individuals receiving the 2023-24 trivalent Northern Hemisphere influenza vaccine. Serum neutralizing antibody titers **(B)** at the time of vaccination and **(C)** 28 days post vaccination against the 2023-24 B/Victoria vaccine strain (B/Austria/1359417/2021 subclade C) and a representative parental (B/Baltimore/0546/2023 subclade C.5) or 2023-24 circulating C.5.1 viruses isolated in the season early (B/Baltimore/JH-663/2023 subclade C.5-E1, B/Baltimore/JH-665/2023subclade C.5-E2) or late (B/Baltimore/JH-547/2024subclade C.5-L). One-way ANOVA with Tukey post-hoc. N = 77. The number of individuals with neutralizing antibody titers ≤ LOD are represented below each dataset **(D)** Fold-change of serum neutralizing antibody titers post vaccination show similar seroconversion rates between all 5 viruses. Bars represent geometric mean and standard deviations. The number of seroconverted individuals (Fold change ≥ 4) is annotated over each dataset. **E)** Patient-paired measurements of neutralizing antibody titers pre (Day 0) and post (Day 28) vaccination. Average fold change between pre- and post- vaccination are shown above each dataset. Two-way ANOVA with Tukey post-hoc. **(F-G)** Total number of tested sera which were below the limit of detection either pre- or post-vaccination.

### The 2023-24 C.5.1 subclade shows increased *in vitro* fitness in later time points compared with the parental C.5 subclade in a temperature-dependent manner

Influenza B viruses belonging to the C subclade continued to circulate over the subsequent two seasons; however, during 2023–24, a late-season surge in cases was associated with viruses harboring multiple substitutions in the HA 120 loop and 190 helix—structurally flexible regions that are critical for receptor binding. The replication of the panel of C.5-related viruses was determined on hNEC cultures. At both 33°C (**Figure 4A**) and 37°C (**Figure 4D**), all the viruses showed comparable replication, though the C.5.1 isolates appeared to have better replication at late times post infection. By determining pre- and post-peak AUC values, it was clear that while the isolates showed variable total virus production pre-peak at both temperatures (**Figure 4B** and **E**), the three newly emerged C.5.1 variants all showed greater virus production at both temperatures post-peak (**Figure 4C** and **F**) compared to the parental C.5 isolate. When temperature was compared across a specific virus (Figure 4 G-J), all the C.5 subclades showed late surges in infectious virus production at 33°C when compared to 37°C These data suggest that circulating C.5.1 viruses have a slower and more gradual increase in virus production prior to peak, but outpace C.5 virus production at late times post infection at both 33°C and 37°C.

**Figure 4.**
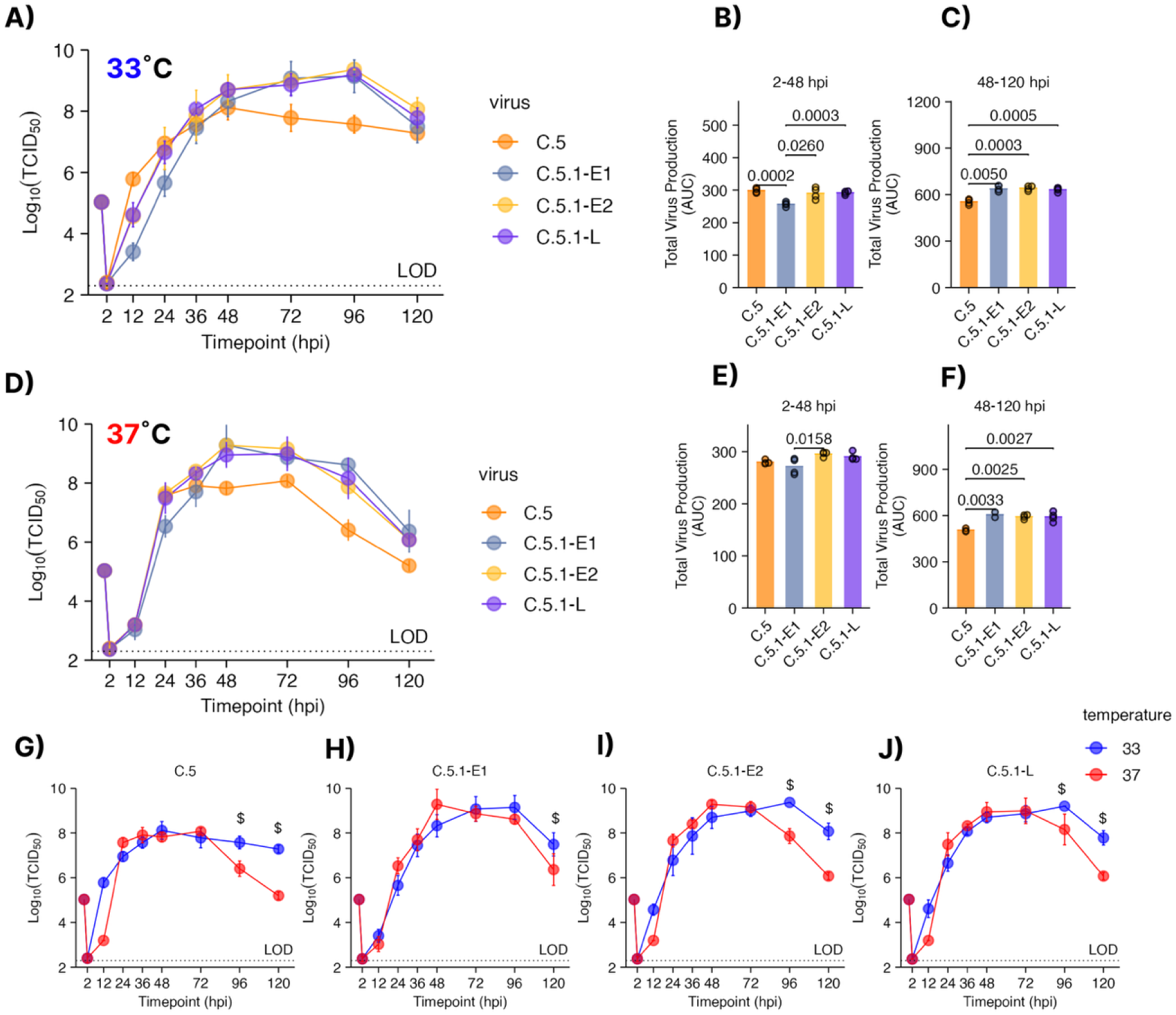
Characterization of Influenza B growth kinetics following the V1A.3a.2 C.5 and C.5.1 subclade emergence at 33°C and 37°C. Low MOI Growth curves on human nasal epithelial cell (hNEC) cultures (MOI = 0.01) at 33°C **(A)** or 37°C **(D)** reveal significantly different kinetics in early and late time points. (B-C) Virus production at 33°C between **(B)** 2-48 hours post infection(hpi) or **(C)** 48-120 hpi. Virus production between at 37°C at **(E)** 2-48 hours post infection (hpi) or **(F)** 48-120 hpi. **(G-J)** Growth curves plotted by virus at 33°C and 37°C per panel. Statistical testing was performed by one way (AUC) or two-way (growth curves) ANOVA with Tukey post host test. Each graph combines two or three experiments with four hNEC wells per experiment.

### Both C.5.1 and C.5 viruses infect multiple cell types in the nasal respiratory epithelium but predominate in ciliated cells

Influenza B viruses have been shown to exhibit both strain and lineage-specific differences in human nasal epithelial cell (hNEC) tropism (13,30,31). Given that circulating C.5.1 viruses demonstrate both increased and prolonged viral production beyond the 48-hour post-infection (hpi) peak observed with the parental C.5 strain, we hypothesized that C.5.1 viruses may exhibit altered or expanded cell tropism in hNEC cultures. We used an established flow cytometry antibody panel to target ciliated (Beta-Tubulin^hi)^, goblet (MUC5AC+) and basal cell (NGFR+) populations. Infected hNECs were gated to exclude debris, and cell doublets followed by identification of infected cells based on positive staining for IBV HA (**Figure 5A-B**) as previously described (13). Because significant differences in hNEC virus production between subclades existed primarily after 48 hpi, we chose 48hpi and 96hpi to assess differences in viral tropism. All 4 contemporary C.5 and C.5.1 viruses infected equivalent numbers of total cells at each timepoint, with more IBV+ cells at 96hpi than 48hpi (**Figure 5C**). Overall, no major differences in infected cell numbers were detected between isolates or within cell populations at 48hpi (**Figure 5D**) and 96hpi (**Figure 5E**). The majority of infected cells were Beta-Tubulin^hi^ ciliated cells followed by MUC5AC+ and NGFR+ cells. Within cell-types, no significant differences were observed between Beta-Tubulin^hi^ and MUC5AC+ cell populations with the exception of C.5.1-E2 across both markers and C.5.1-E1 for MUC5AC+ alone (**Figure 5F-H**). No significant differences in infection levels were observed among NGFR-positive (basal) cell populations across viruses. These findings suggest that enhanced infectious virus production, rather than differences in cell tropism or numbers of infected cells, drives the prolonged viral replication observed in C.5.1 viruses.

**Figure 5.**
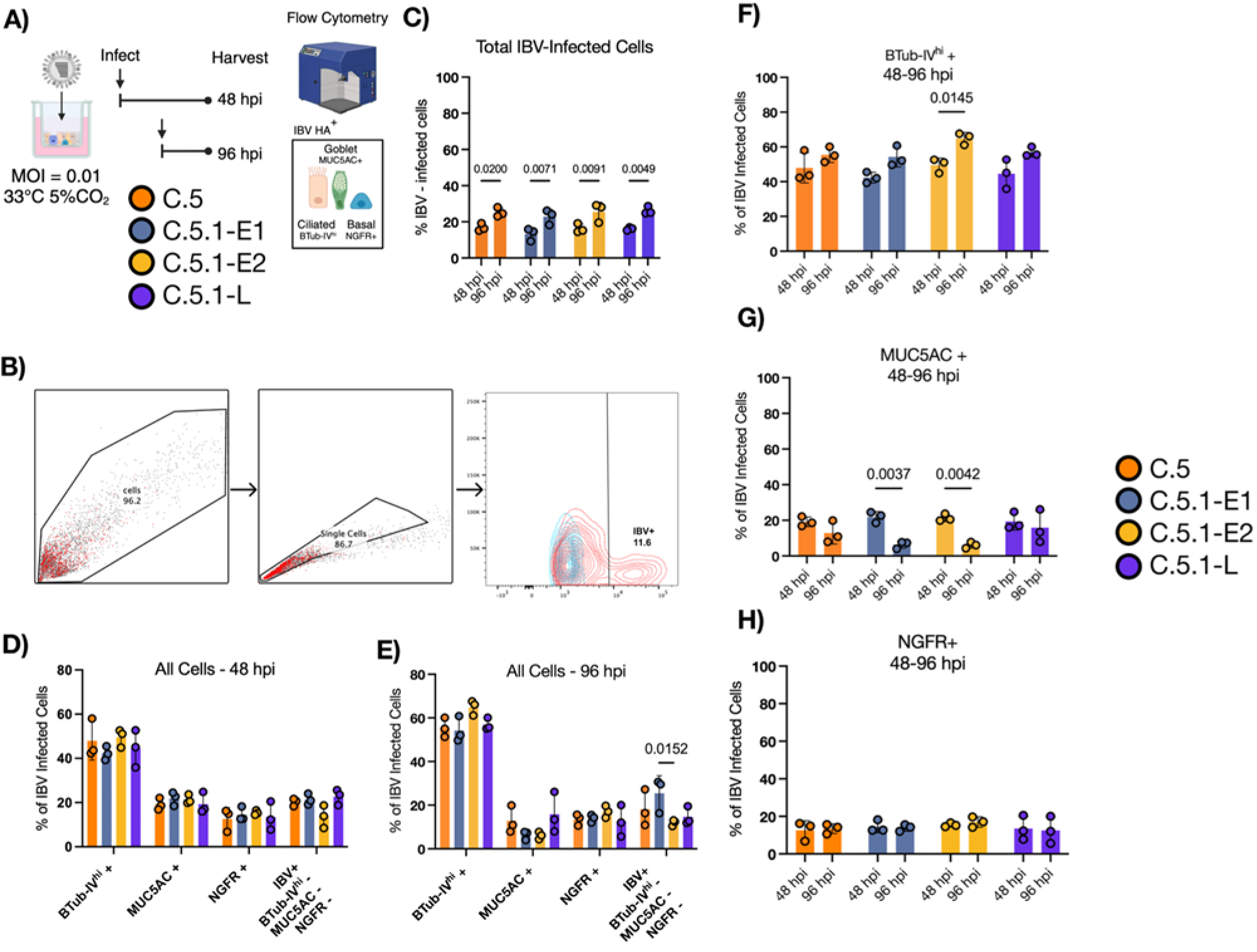
C.5 parental and 2023-24 C.5.1 Influenza B viruses infect human nasal epithelial cell subsets with similar breadth. **(A-B)** Experimental design and gating strategy of IBV-infected hNECs via flow cytometry. Cells were gated by removing debris and doublet cells. Human nasal epithelial cell markers were used to assess cell tropism including BTub-IV as a marker of mature ciliated cells, MUC5AC as a marker of mucin-producing cells and NGFR (CD271) as a marker of respiratory basal cells. Data represents 48- and 96-hour endpoint infections for two independent runs and 3 replicates per virus. **C)** Percentage of total IBV-infected cells. IBV-infected hNECs were identified using HA monoclonal antibodies for IBV HA. **(D-E)** The specific cell types infected over the course of infection were identified using antibodies specific for the same cellular targets at 48 and 96 hours, respectively. (**F**) Percent of IBV-infected ciliated cells infected at 48 and 96 hours. (**G**) Percent of IBV-infected MUC5AC + cells at 48 and 96 hours. (**H**) Percent of IBV-infected NGFR+ cells at 48 and 96 hours. Percentages of IBV-infected hNECs were statistically compared between two groups using multiple *t*-tests.

### The prevalence of the C.5.1 subclade is geographically biased toward the Americas

IBV has historically been observed to be geographically limited prior to broad epidemic spread (32). Using 1655 down sampled human IBV specimen sequences from GISAID, we assessed the phylogeographic distribution of the parental C.5 (**Figure 6B**) and circulating C.5.1, C.5.6 and C.5.7 subclades isolated between January 2020 and January 2024 (**Figure 6B**). The parental C.5 subclade was not biased by geographical origin, however, C.5.1 had biased prevalence towards North, South and Central America compared to European countries (**Figure 6B**). Inversely, C.5.7 was sequenced very infrequently in the Americas and primarily found in European and Asian countries during the NH2023-24 season (**Figure 6D**). C.5.6 showed no clear geographic bias by our analysis (**Figure 6E**).

**Figure 6.**
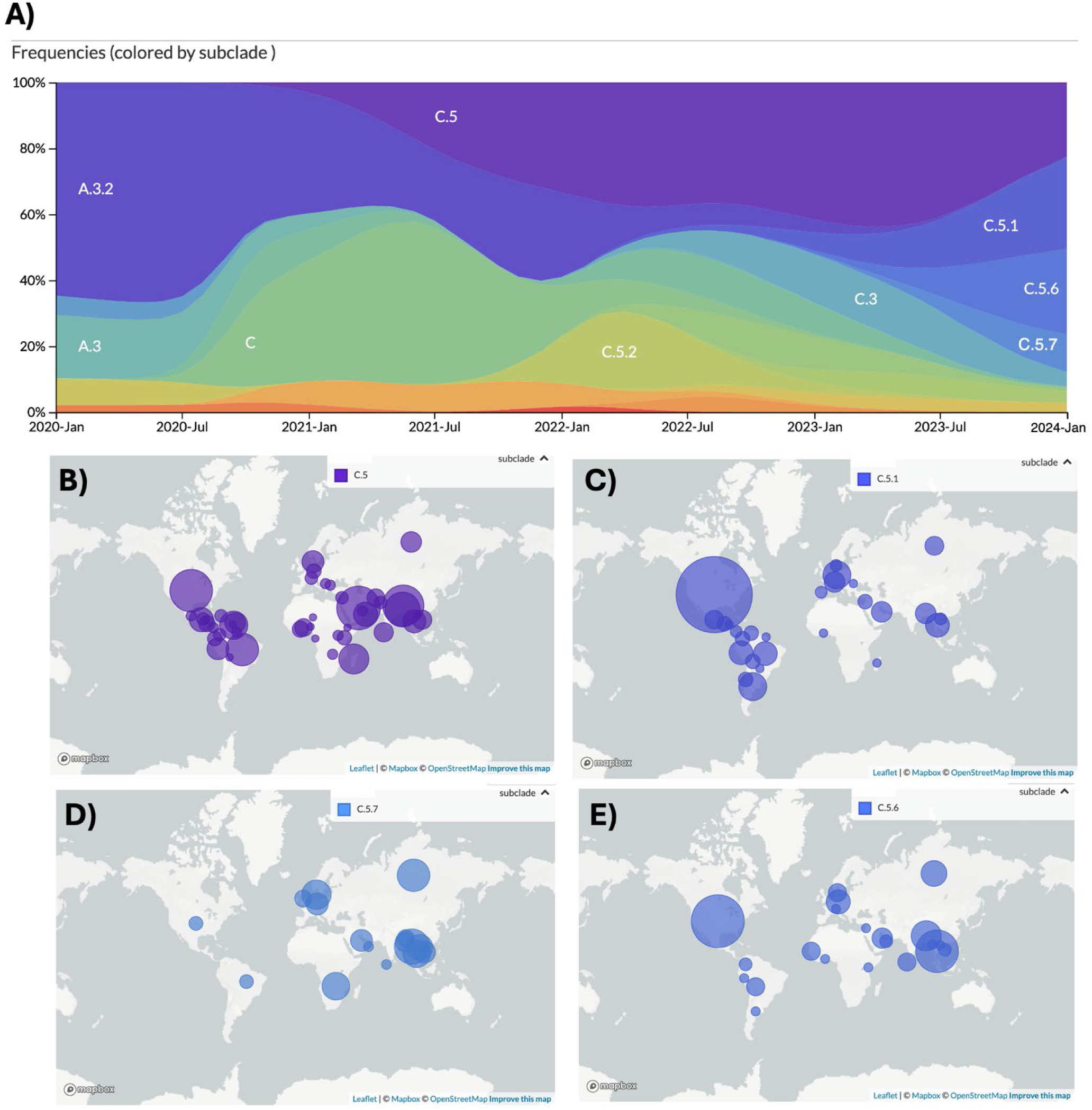
2023-25 C.5.1 subclade predominance is biased to the Americas. (A) Global IBV subclade frequency between January 2020 and January 2024 of 1655 genomes down sampled by collection week and country of isolation as reported to GISAID. Bubble plots representing relative abundance by country are shown for the parental (B) C.5, (C) C.5.1, (D) C.5.7, and (E) C.5.6. Bubble size represents relative frequency of each respective subclade.

## Discussion

The accumulation of mutations in circulating seasonal influenza viruses leads to antigenic drift, necessitating annual vaccine updates and influencing viral fitness (1,33). Selective pressures act on Influenza viruses at a global scale due to widespread vaccination, natural infection, and international travel, all of which shape viral antigenic profiles and transmission dynamics (18,34,35). Furthermore, mutations can additionally impact various aspects of viral fitness including replication kinetics (9,36,37), budding (12) and receptor binding (9,24,38).

Since the disappearance of the B/Yamagata lineage in early 2020, B/Victoria lineage viruses have resurged (3,26) and has been marked by rapid evolutionary changes within the genome that diverge from historical patterns of influenza B virus evolution. While B/Victoria viruses do undergo antigenic drift, the rate is substantially slower than that of influenza A viruses (39). Historically, B/Victoria lineages have exhibited localized circulation, often persisting within regional populations before expanding into globally fixed clades (32).

The findings presented in this manuscript reveal critical insights into the evolutionary trajectory of influenza B viruses during the 2023-24 season, particularly regarding the emergence and fitness characteristics of the C.5.1 subclade. To our knowledge, this study is the first to assess simultaneous in vitro replication, antigenic drift and primary cell tropism of Influenza B viruses collected in the wake of C. 5. 1. subclade emergence. In the post-COVID19 era, B/Victoria viruses have re-emerged with increasing prevalence, fixed to 1 clade, V1A.3a.2 globally. While mutations in individual influenza proteins can affect viral replication and antigenicity, studies specifically investigating the impact of individual mutations in influenza B are limited and often focus only on clade-level differences or specific HA antigenic sites (23,40). Our surveillance data reveal that this resurgence was antigenically static, at least with respect to serum neutralizing antibody titers. Clade C.5 has nearly fixed globally diversifying into several subclades with recurrent HA1:197E and HA1:183K substitutions, both within the RBS and 190 helix (17,20,23). The 2023-24 season in Baltimore, MD was driven by 2 waves of cases which were genomically distinguishable through phylogenomic reconstruction at each genomic segment. As of 2024, no single IBV subclade has fixed globally with C.5.1 rapidly increasing across North America and C.5.7 increasing with high frequency in China (**Figure 5**) (41).

We sought to understand how these mutations, at the subclade level, affect antigenicity relative to the 2023–24 vaccine strain and parental C.5 viruses. Using infectious virus neutralization assays and a serum panel from vaccinated healthcare workers, we found that sera collected at baseline and 28 days post-vaccination neutralized C.5 and C.5.1 viruses equally well compared to the vaccine strain.

Our findings suggest no evidence of antigenic drift between the parental C virus and circulating C.5 and C.5.1 viruses isolated in Baltimore, Maryland (Figure 2). Fold-increase in neutralizing titers at day 28 following was similar for all 5 viruses. It is important to note that healthcare workers represent a highly vaccinated population, with mandatory vaccination leading to over 90% of the individuals in the cohort having received the influenza vaccine annually for the past three years (42). This result was surprising given that residue 197 in the HA protein is a known site of positive selection and has previously been associated with antigenic drift. Notably, our in vitro replication data demonstrate that C.5.1 viruses sustain higher viral output at later time points in primary human nasal epithelial cells, indicating a shift in fitness kinetics (**Figure 3**). Historical viruses circulating prior to the introduction of V1A.3a.2 (subclade C) were also included to assess greater longitudinal changes in fitness. Isolation and assessment of 2 strains from the V1A.3 clade revealed similar kinetics to contemporary viruses suggesting that C.5 may be a transitionary strain which was selected for during the low-circulation of IBV following COVID-19 and up to the 2022-23 season (5).

Temperature has been demonstrated to impact viral replication kinetics and the epithelial host transcriptional landscape in the context of Influenza A and SARS-CoV-2 infection (27,43). Takada et al., 2024 recently described that Influenza B viruses may be more sensitive to higher temperatures at 37°C compared to cooler ambient temperatures present in the nasal epithelium than Influenza A viruses (44). Recently circulating C.5.1s have a more dramatic attenuation at 12hpi compared to historical A.3.2 and C strains except for the strain-specific attenuation of strain A.3.2-1 (**Figure 2**). Our works supports only a strain-strain specific attenuation for 3 of our 4 contemporary C.5.1 Influenza B viruses assess for fitness at 37°C compared to 33°C which is especially apparent at 12-hour post infection (**Figure 3I-L**). A possible explanation for this phenomenon could relate to HA’s temperature-dependent acid stability at 33°C further supporting the idea that IBV prefers the temperature of upper airways (45).

Mutations within and surrounding the RBS of IBV HA have been shown to alter receptor binding breadth between a2,3 and a2,6 sialic acid receptors (20,21,46,47). All late C.5.1 viruses, which rapidly replaced early C.5.1 strains, harbor the A202V RBS flanking mutation which sits within the 190 helix of the RBS (21). Goblet cells primarily express a2.3 terminally sialylated glycans as opposed multi-ciliated cells in the upper airway which primarily express a2,6 within the respiratory tract (48,49). All strains, including C.5 and early/late C.5.1 viruses primary infect Beta-Tubulin^hi^ ciliated cells over goblet and basal cell populations, a finding previously demonstrated in influenza infected hNEC cultures (13,30).

However, MUC5AC + infected IBV cells infected with C.5.1 early strains dropped significantly between 48 and 96 hpi where the parental and C.5.1-L strains did remain similarly infected. These findings suggest that extended viral replication observed in C.5.1 strains are unlikely to be driven by changes in epithelial cell-type targeting but rather enhanced infectious virus production at later time points.

Finally, we describe the relative distribution of the C.5 parental subclade and the circulating C.5.1, C.5.6, and C.5.7 subclades, which account for the majority of IBV cases sequenced and submitted to GISAID during the 2023–24 season globally, including those from Baltimore. C.5.6 is defined by a substitution to asparagine at position 129, while C.5.7 is marked by a glycine at position 128 (50). Both of these residues are in critical regions of the 120-helix of the HA1 head domain(51). Notably the adjacent site 127 has previously been implicated in antigenic drift and may have played a role in the divergence of the two major influenza B lineages. Consistent with historical patterns, B/Victoria viruses continue to exhibit regionally constrained diversification, with subclades often circulating within fixed geographic boundaries for extended periods before achieving broader global spread (7,52,53,53,54). This geographic isolation facilitates local adaptation, including the accumulation of substitutions at key antigenic sites. As IBV cases continue to undergo interregional transmission and reassortment opportunities expand in the post-pandemic era, previously confined subclades may re-enter global circulation, raising the risk of antigenically novel viruses escaping existing immunity (55). Ongoing surveillance and functional characterization of these geographically structured lineages remain critical for anticipating future antigenic evolution and guiding vaccine strain selection.

## Materials and Methods

### Ethics statement and human subjects

Serum used in this study was obtained from healthcare workers recruited during the annual Johns Hopkins Hospital employee influenza vaccination campaign in the Fall of 2023 by the Johns Hopkins Center for Influenza Research and Response (JH-CEIRR). Pre- and post-vaccination (∼28 day) human serum were collected from subjects, who provided written informed consent prior to participation. The JHU School of Medicine Institutional Review Board approved this study, IRB00288258. Virus was isolated for this study from deidentified influenza A virus H1N1 positive samples, collected from patients who provided written informed consent during the 2024-25 influenza season at the Johns Hopkins Hospital, under the JHU School of Medicine Institutional Review Board approved protocol, IRB00091667.

### Cell Culture

Madin-Darby canine kidney (MDCK) cells (provided by Dr. Robert A. Lamb, Northwestern University) and MDCK-SIAT cells (provided by Dr. Scott Hensley, University of Pennsylvania) were maintained in complete medium (CM) consisting of Dulbecco’s Modified Eagle Medium (DMEM) supplemented with 10% fetal bovine serum, 100 units/ml penicillin/streptomycin (Life Technologies) and 2 mM Glutamax (Gibco) at 37 °C and 5% CO2. Human nasal epithelial cells (hNEC) (PromoCell) were cultivated as previously described.

### Viral RNA Extraction and Whole Genome Amplification

After amplification, concentrations were checked using the Qubit 2.0 fluorometer (ThermoFisher Scientific). 2μL of PCR product was prepared for sequencing using the native barcoding genomic DNA kit (EXP-NBD196) and the NEBNext ARTIC Library Prep kit, following the manufacturer’s instructions (New England BioLabs / Oxford Nanopore Technologies). 50ng of the barcoded library was sequenced using the R9.4.1 flow cell on a GridION (Oxford Nanopore Technologies). Resulting fastq files were demultiplexed using the artic_guppyplex tool (Artic version 1.2.2). Nucleotide sequence assembly was performed using the FLU module of the Iterative Refinement Meta-Assembler (IRMA version 1.0.2), using IRMA’s default settings, which include a minimum average quality score of 24 and a site depth of 100.

### Phylogenetic Analysis of Influenza B Segments and Concatenated Genomes

Consensuses sequences of segments 1-8 were pipped through custom scripts for ingestion, phylogenetic reconstruction and visualization. Briefly, consensus FASTA segment sequences were called for type (Influenza A vs Influenza B) and subtype or lineage (H1N1, H3N2, or Victoria) using BLAST+ and custom python scripts available at https://github.com/Pekosz-Lab/NH20-24_influenzab/blob/main/flusort/flusort.py. Typed and subtyped sequences from Johns Hopkins Hospital along with vaccine reference sequences B/Austria/1359417/2021 and B/Washington/02/2019 were passed through the fludb (https://github.com/Pekosz-Lab/NH20-24_influenzab/tree/main/fludb) for standardization. The download.py script was used to separate segment sequences and metadata into respective directories and concatenate genomes. Influenza B clade, subclade, HA/NA glycosylation, alignment coverage and overall quality status was called by Nextclade CLI. Time-resolved phylogenies were constructed using Augur v26.0.0 (56). All scripts and workflows used for sequence ingestion, processing, phylogenetic inference, and visualization are available in the project repository (https://github.com/Pekosz-Lab/NH20-24_influenzab).

### Virus Isolation on Human Nasal Epithelial Cells *(hNECs)*

Viral stock preparation was performed as described previously described using nasopharyngeal swabs from Influenza B-positive individuals during the 2023–2024 Northern Hemisphere influenza season at Johns Hopkins Hospital (2).

### Human Nasal Epithelial Cell (hNEC) growth curves

hNEC growth curves were performed on fully differentiated hNECs grown at an air–liquid interface. Virus inoculum was made by diluting virus stocks to an MOI of 0.01 in hNEC infection media. Wells were washed three times with hNEC infection media and inoculum was added to the wells. Inoculum was incubated on the cells for two hours, after which it was removed, and cells were washed three times with PBS and left at an air–liquid interface. For each timepoint, 100 µl of hNEC infection media was added to the wells and incubated at 33°C for 10 min. Media was then collected and cells were left at ALI. Virus concentration in collected media was determined through TCID50.

### Serum Neutralization Assay

Serum samples used for this analysis originated from Johns Hopkins Medical Institute healthcare workers recruited from the Johns Hopkins Center of Excellence for Influenza Research and Response (JH-CEIRR). Recruitment occurred during the annual employee influenza vaccination campaign in the Fall of 2019. Participants were vaccinated with split inactivated quadrivalent vaccine formulations. Influenza vaccination is required of all Health Care Workers at Johns Hopkins University and, therefore, most of the study participants have had an influenza vaccine over the past three years. Pre-vaccine and approximately 28-day post-vaccination samples were used for analysis. Subjects provided written informed consent prior to participation. The JHU School of Medicine Institutional Review Board approved this study, IRB00288258. Serum samples were first treated (1:3 ratio serum to enzyme) with Receptor Destroying Enzyme (Denka-Seiken, Tokyo, Japan) and incubated overnight at 37 °C followed by inactivation at 57 °C for 35 min. Serum was diluted 2-fold in IM (Dulbecco modified Eagle medium (Sigma), with 10% penicillin/streptomycin (Gibco), 10% L-glutamine (Gibco), 0.5% BSA (Sigma), and 5 µg/mL of N-acetyl trypsin (Sigma) at 37 °C and 5% CO_2_) and 100 TCID50 was added for a one-hour incubation at room temperature. Serum Sample/Virus was used to infect a confluent layer of MDCK-SIAT-1 cells. The inoculums were removed after 24 hours, IM supplemented with 2.5µg/mL N-acetyl Trypsin was replaced, and cells were incubated for 96 hours at 37°C. Plates were fixed and stained as described previously. The Neutralizing Antibody titer was calculated using the highest serum dilution that led to greater than 50% CPE. Statistical analysis was completed comparing means of pre- and post-neutralizing responses as well as between lineages. Mean differences were calculated and Sidak’s multiple comparisons test was used to assess significant differences. Statistical differences were set at *p* ≤ 0.05.

### Flow Cytometry of Human Nasal Epithelial Cells

For the 48- and 96-hour time point experiments, hNEC cultures were infected with IBV clinical isolates at an MOI of 0.01. Cells from both infected samples and uninfected controls were harvested, creating a single-cell suspension after a 30-minute incubation in 1X TrypLE. The cells were then resuspended in trypsin stop solution, washed, and resuspended in 1X PBS. All washes were carried out between blocking, primary, secondary or conjugated antibody staining using either 1X PBS or BD Perm/Wash Buffer (after Fixation/Permeabilization) at a centrifuge speed of 400× *g* at 4 °C. Samples and controls were stained with AQUA viability dye (1 μL/1 × 10^6^ cells) for 30 min at room temperature (RT). Following staining, cells were washed and resuspended in BD Fixation/Permeabilization solution, and they were incubated for a minimum of 30 min at 4 °C. Cells were blocked with 7% normal goat serum in BD Perm/Wash Buffer for 1 hour at 4 °C. Next, cells were incubated with primary antibodies and diluted in BD Perm/Wash buffer at appropriate concentrations for one hour at RT. Subsequently, cells were incubated with secondary antibodies, as described for the primary antibodies, for 30 min at RT. After secondary antibody staining, the same procedure was repeated for incubation with conjugated antibodies for 30 min at RT. Cells were finally resuspended in FACS Buffer and filtered through a 35 μM strainer cap into FACS tubes.

Stained suspensions were analyzed using a BD Symphony Flow Cytometer with DIVA software v7.0. The following laser-emission filter-(fluorochrome groupings) were used: 488 nm-530/30-(AF488,PE), 633 nm-660/20-(AF647), 405 nm-525/50-(AQUA/BV605). Controls included single stained cells, fluorescence minus one control for gating assistance, and uninfected IBV HA stained controls to ensure absence of non-specific staining. Data analysis was conducted using FlowJo v10. The gating strategy consisted of excluding debris, selecting single cells, and identifying influenza B HA positive cells.

### Tissue Culture Infectious Dose 50 (TCID_50_) Assay

MDCK-SIAT-1 cells were seeded in a 96-well plate 2 days before assay and grown to 100% confluence. Cells were washed twice with PBS+ then 180 µL of IM was added to each well. Ten-fold serial dilutions of virus from 10^−1^ to 10^−7^ were created and then 20 µL of the virus dilution was added to the MDCK-SIAT-1 cells. Cells were incubated for 6 days at 33 °C then fixed with 2% formaldehyde. After fixing, cells were stained with Naphthol Blue Black, washed and virus titer was calculated using the Reed and Muench method

### Virus Seed and Working Stocks

The earliest clinical isolate sample that showed presence of infectious virus was selected. A T75 flask of confluent MDCK-SIAT-1 cells was infected at an MOI of 0.01. Working stocks for each clinical isolate were generated by infecting a T75 flask of MDCK-SIAT-1 cells at an MOI of 0.001 for one hour at room temperature while rocking. The inoculum was removed, and cells were placed in a 33 °C incubator and monitored daily for CPE. Working stock was harvested between 3 and 5 days later, when CPE was seen in 75–80% of the cells. Harvested media were centrifuged at 400× *g* for 10 min at 8 °C to remove cell debris, and the resulting supernatant was aliquoted into 500 µL and stored at −80 °C infectious virus quantity of working stocks was determined using TCID-50 assay. Seed and working stocks of the egg-adapted vaccine strains of IBV (see **Table 2**) were grown directly in MDCK-SIAT-1 cells as described above. IBV vaccine strains were kindly provided by Johns Steel, Centers for Disease Control (CDC).

### Cellulose Plaque Assays

MDCK cells were grown in complete medium to 100% confluency in 6-well plates. Complete medium was removed; cells were washed twice with PBS containing 100 µg/mL calcium and 100 µg/mL magnesium (PBS+) and 250 µL of inoculum was added. Virus dilution was conducted by serially diluting the virus stock 10-fold each time until 10^−6^. Cells were incubated at 33 °C for 1 h with rocking every 10 min. After 1 h, the virus inoculum was removed and phenol-red free MEM supplemented with 3% BSA (Sigma), 100 U/mL of penicillin and 100 μg/mL of streptomycin mixture (Life Technologies), 2 mM Glutamax (Gibco), and 5 µg/mL N-acetyl trypsin (Sigma), 5 mM HEPES buffer and 1% cellulose was added. Cells were incubated at 33 °C for either 48 or 72 hours and then fixed with 4% formaldehyde. Cells were stained with Naphthol Blue Black. Plaque size was analyzed in Fiji.

### Statistical analyses

All statistical analyses were performed in GraphPad Prism 10.4.2 or Rv4.5.1. Growth curves were analyzed using 2-way ANOVA with Tukey post-hoc test. Area under the curve (AUC) was calculated for each growth curve replicate and AUC values were compared using one-way ANOVA for differences in total virus production. Plaque assays were analyzed using Kruskal–Wallis ANOVA with Dunn’s multiple comparison test. Serology was analyzed using either 2-way ANOVA or 1-way ANOVA, both with Tukey post-hoc test.

## Data availability

The datasets used and/or analyzed during the current study are available through the Johns Hopkins Research Data Repository at **[INSERT DOI WHEN RELEASED].** An interactive build of the concatenated genome and each of the 8 individual segments is available at https://nextstrain.org/groups/PekoszLab-Public/akine/ibv2025/genome. All scripts generated in this manuscript are available at https://github.com/Pekosz-Lab/NH20-24_influenzab

## Acknowledgments

This work was supported by National Institutes of Health (NIH) contracts N272201400007C and N7503021C00045 for the Johns Hopkins Center of Excellence for Influenza Research and Response, NIH T32 AI007417 and Richard Eliasberg Family Foundation. The authors thank the healthcare workers who enrolled and participated in the Johns Hopkins Center for Excellence in Influenza Research and Surveillance study. We are grateful for the efforts of the clinical coordination team at JHH who collected samples. We thank the laboratories of Heba Mostafa, Kimberly Davis, Sabra Klein, and Andrew Pekosz for discussion of data and future directions

