## Supplementary material for "*In-vitro* characterization of 2019-24 Influenza B Viruses reveals increased temperature-dependent fitness in later timepoints independent of antigenic drift": suppl figures 1 and 2

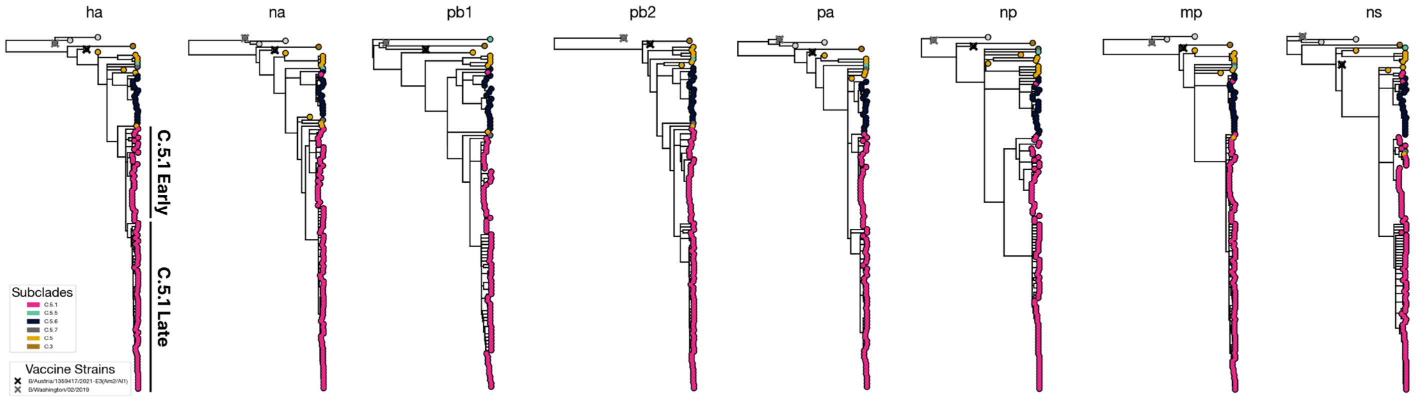


**Figure S1.** Phylogenetic trees of segment genomes with sequencing date-resolved internal branches. Tree-tips are colored by HA subclade designation or vaccine identity. An interactive build of the concatenated genome and each of the 8 individual segments is available at https://nextstrain.org/groups/PekoszLab/akine/ibv2025/genome

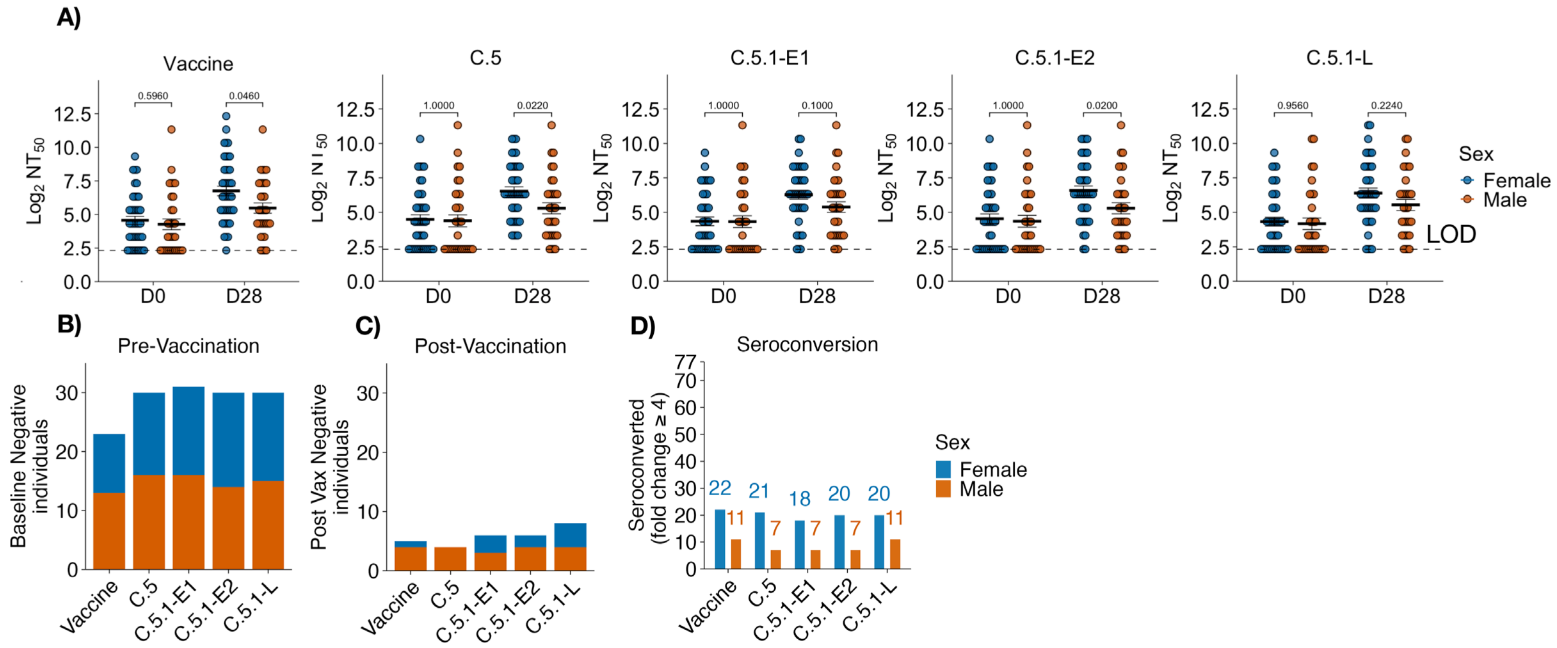


**Figure S2. Serum neutralizing antibody titers ($NT_{50}$) stratified by sex in serum of healthcare workers (A)** Serum neutralizing antibody titers against the 2023-24 B/Victoria vaccine strain (B/Austria/1359417/2021 subclade C and a representative C.5 parental (B/Baltimore/0546/2023) or 2023-24 circulating C.5.1 viruses isolated in the season early against the 2023-24 B/Victoria vaccine strain (B/Austria/1359417/2021 subclade C) and a representative parental (B/Baltimore/0546/2023 subclade C.5) or 2023-24 circulating C.5.1 viruses isolated in the season early (B/Baltimore/JH-663/2023 subclade C.5-E1, B/Baltimore/JH-665/2023subclade C.5-E2) or late (B/Baltimore/JH-547/2024subclade C.5-L). Post vaccination neutralizing antibody titers were significantly higher in females against the vaccine strain, C.5 and C.5.1-E2 while C.5.1-E1 and C.5.1-L do not reach statistical significance. Wilcox-test with Bonferroni post-hoc correction. N = 77. Stacked bar plot filled by male or females for where the individual's sera was below the NT50 limit of detection (NT50 ≤ 10) for pre **(B)** and post **(C)** vaccination. **(D)** Seroconversion (post vaccination NT50 fold change ≥4) stratified by males vs females.
